# Cohort Profile: Baseline characteristics and design of the McMaster Monitoring My Mobility (MacM3) Study, a prospective digital mobility cohort of community-dwelling older Canadians from Southern Ontario

**DOI:** 10.1101/2025.05.14.25327626

**Authors:** Marla K Beauchamp, Renata Kirkwood, Cody Cooper, William McIlroy, Karen Van Ooteghem, Kit Beyer, Julie Richardson, Ayse Kuspinar, Paul D. McNicholas, K. Bruce Newbold, Darren M. Scott, Parminder Raina, Qiyin Fang, Paula Gardner, Manaf Zargoush, Jinhui Ma, Sachi O’Hoski, Talha Rafiq, the MacM3 investigators

## Abstract

**Purpose:** The McMaster Monitoring My Mobility (MacM3) study aims to understand trajectories of mobility decline in later life using multi-sensor wearable technology. To our knowledge, MacM3 is the first major cohort to combine accelerometry and GPS to track real-world mobility in community-dwelling older adults.

**Participants:** Between May 2022 and May 2024, MacM3 recruited 1,555 community-dwelling older adults (mean age 73.9 years, SD = 5.5) from Hamilton and Toronto, Ontario. Of the cohort, 68.4% were female, 62.4% married/partnered, 75.3% had post-secondary education, and 63.0% had ≥3 comorbidities. Most were Canadian born (69.4%) and White/Caucasian (88.0%), with greater ethnocultural diversity observed at the Toronto site.

**Findings to date:** At baseline, 56.7% of participants reported no mobility limitations, 15.9% had preclinical limitations, and 27.4% had minor mobility limitations. Mean gait speed for the total sample was 1.23 m/s, with a mean Timed Up and Go time of 9.4 seconds and a 5x sit-to-stand time of 13.0 seconds. A total of 1,301 participants had valid wrist-worn device data, and 1,008 participants who agreed to wear the thigh-worn device had valid data (≥7 days with ≥10 hours of wear per day). Step count data (n = 1008) revealed a mean of 8,437 steps per day (SD = 2,943), with 5,073 steps in the lowest quartile and 12,303 steps in the highest.

**Future plans:** Ongoing work aims to develop predictive models of mobility decline by integrating wearable, clinical, and environmental data. Pipeline enhancements will enable GPS/inertial measurement unit fusion to explore mobility-environment interactions and support aging-in-place tools.

## INTRODUCTION

In the 2015 World Report on Ageing and Health, the World Health Organization defines healthy aging as the ‘process of developing and maintaining the functional ability that enables well-being in older age’.^1^ Central to this concept is mobility — the ability to move, walk, and use transportation within our homes and communities.^2^ Mobility is a cornerstone of healthy aging, reported to be of utmost importance to older people as they age, and has been labeled by many aging experts as the ‘6th vital sign’.^3–5^ With population aging, the number of older adults living with mobility limitations will rise dramatically over the next four decades. In Canada, one in two older adults report reduced mobility compared to the previous decade, with 24-33% facing challenges in conducting daily activities that require physical capabilities, such as climbing stairs and walking.^6–8^ These limitations can signal early neurological, musculoskeletal, or cardiorespiratory health changes and predict adverse outcomes like falls, hospitalization, institutionalization, and mortality.^9–12^

Despite ongoing efforts to understand predictors and trajectories of mobility decline in older adulthood, knowledge gaps remain, particularly in understanding the varied trajectories of mobility changes across different age groups and functional levels.^13–18^ Previous studies have predominantly focused on older populations (age 70+) already experiencing mobility limitations or relied on self-reported measures of function, which may not accurately capture day-to-day mobility challenges.^13,14,16–18^ As such, our understanding of the actual level and types of mobility undertaken by individuals in their daily lives is limited.^19^ By expanding our research to include a broader age range and incorporating device-based measures of real-world mobility, we can effectively identify early signs of decline and inform interventions to delay or prevent the onset of disability.

Advancements in wearable technology have revolutionized our ability to continuously monitor unsupervised mobility in individuals’ daily lives, both at home and in the community (‘free-living’).^20–22^ This innovation has generated substantial interest and great potential to utilize wearables for prevention, health management, and personalized care, especially among older adults who increasingly recognize the value of self-monitoring their mobility as part of their health regimen.^23,24^ Our consultations with Canadian stakeholders (i.e., older adults, caregivers, and industry and community partners) confirmed that older adults have a strong interest in using such technology to monitor their mobility and enhance their health. Given that mobility data obtained using wearable technology (i.e., digital mobility measures) reflect real-world functioning, these measures hold exceptional potential for monitoring and predicting health outcomes and promoting aging-in-place.^25,26^

In 2022, we established the McMaster Monitoring My Mobility (MacM3) research platform, with the primary objective of characterizing later-life trajectories of early mobility decline through longitudinal, multi-sensor-based data collected from a large cohort of older adults exhibiting differing levels of mobility at baseline. This pioneering initiative represents the first major longitudinal study to evaluate real-world mobility with frequent repeated measures in community dwelling older adults using a wearable system that integrates GPS (Global Positioning Systems) and accelerometry.^27^ Our methodology centers on a wrist-worn device that synchronizes time-based accelerometry with GPS data, complemented by a thigh-worn accelerometer. The unique combination of accelerometry and geolocation data provides a holistic view of mobility, capturing interactions between individuals and their environments. Such a comprehensive approach enhances our capacity to detect early signs of mobility decline and develop strategies to mitigate mobility-related adverse health impacts in older adulthood.

## COHORT DESCRIPTION

### Study design and recruitment

MacM3 is a prospective cohort study with 24 months of follow-up initiated in May 2022 at the Hamilton, Ontario site and expanded to the Dixon Hall social services site in Toronto in January 2023. The study targeted individuals aged 65 years and older residing within a 50 km radius of McMaster University in Hamilton or Dixon Hall in Toronto. All participants provided verbal informed consent. Eligibility criteria included proficient in English or either English or Mandarin (at Dixon Hall) and classification into one of three mobility groups based on the concept of preclinical mobility limitation^28^ measured by the Preclinical Mobility Limitation classification (PCML)^29^: 1) no mobility limitation, 2) preclinical mobility limitation, or 3) minor mobility limitation. The PCML classification is based on self-reported difficulty with three mobility tasks: walking 0.5 km, walking 2 km, and climbing one flight of stairs. For each task, participants are asked whether they are: (1) able to manage without difficulty, (2) able to manage with some difficulty, (3) able to manage with a great deal of difficulty, (4) able to manage only with the help of another person, or (5) unable to manage even with help. Participants who report being able to manage any task without difficulty (response 1) are further asked about task modification using a set of follow-up questions. These include: “I am able to walk 0.5 km the same way I always have,” “I need to rest in the middle of the walk,” “I walk 0.5 km less often than I used to,” “It takes me longer to walk 0.5 km than it used to,” and “I am more tired after walking 0.5 km than I used to be.” Participants who report no task modification—i.e., they perform all tasks the same way as before—are classified as having **no mobility limitation**. Those who report modification of any task (e.g., reduced frequency, slower performance, fatigue, or need for rest) are classified as having a **preclinical mobility limitation**. Individuals who report being able to manage a task with some difficulty (response 2) are classified as having a minor mobility limitation, while those who report great difficulty, needing help, or being unable to complete a task (responses 3, 4, or 5) are classified as having a **major mobility limitation**. While individuals with major mobility limitation were excluded at baseline, they could be reclassified into this category during follow-up assessments. Additional exclusion criteria included inability to communicate in required languages, significant visual or hearing impairments that could affect safety during assessments and plans to relocate outside the study area within two years.

We engaged in a multi-pronged recruitment strategy aimed at enhancing the diversity of participants in the study (in ethnicity, race, income level, culture, ability, and region), and ensuring the data reflects diverse participant experiences. Our recruitment strategy employed a combination of community-based approaches, including word of mouth, advertisements in local newspapers, and online platforms. Targeted mail campaigns were conducted using Canada Post Snap Admail, focusing on postal routes and codes that matched the study’s demographic in lower-income areas, and advertisements were placed on relevant public transportation routes. Our recruitment strategy was designed to reach adults aged 65 and older—including males, females, individuals of any gender identity, and people living in both higher- and lower-income areas—to ensure a diverse and inclusive sample.

Our second recruitment site, Dixon Hall, serves a large, diverse population, including many individuals who are underrepresented in research, particularly those facing social challenges such as poverty, isolation, and health inequities. Our partnership with Dixon Hall, facilitated by the McMaster Institute for Research on Aging (MIRA) | Dixon Hall Centre, allows us to engage with a population that is often excluded from research studies, ensuring that the data collected are more generalizable to the broader community of older adults. In order to build trust and access, we embedded researchers at the Dixon Hall site, recruiting participants active in social and recreation groups via a snowball method, where participants invited peers to participate. While the Hamilton participants tend to be more homogenous (Caucasian and middle-income), Dixon Hall participants had more diversity in race, income level, ethnicity, and primary spoken language (see Table 1). Additionally, a small subset of participants (*n* = 148) was recruited from the MIRA Intergenerational Study (MIRA-iGeN) platform, which includes 5,000 individuals pre-recruited by Canadian Viewpoint Inc. through phone, online, and mail-out methods.

**Table 1.**
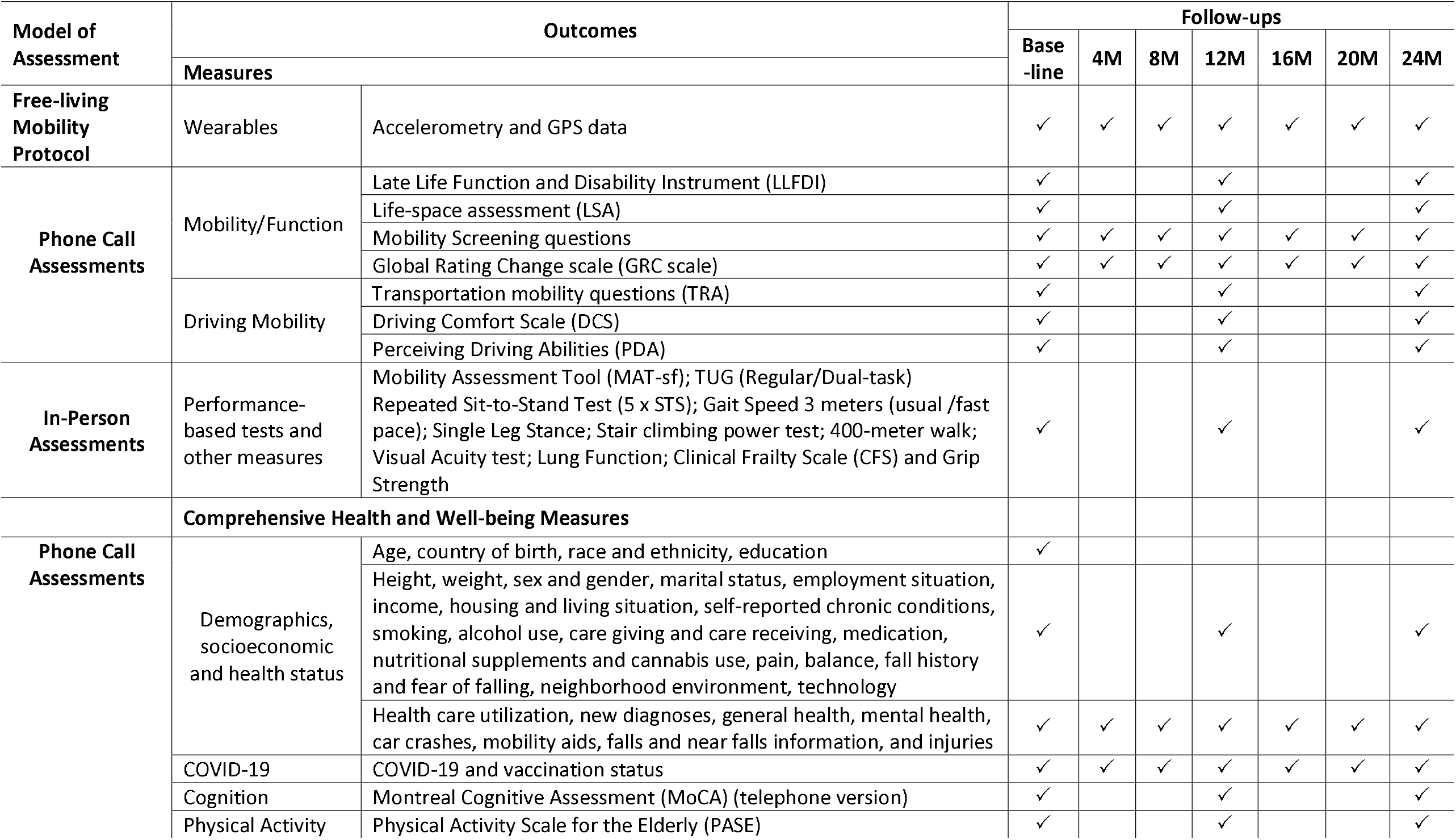
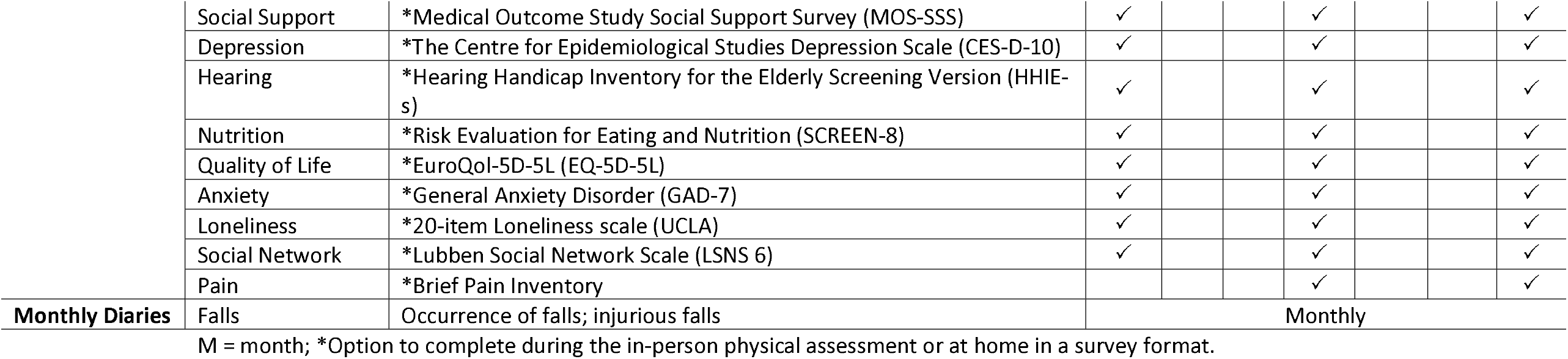
Outcome measures and time points of the MacM3 cohort study.

Our sample size was designed to support MacM3’s main objectives: to compare mobility patterns among older adults with varying levels of early mobility limitation; identify distinct mobility trajectories and their characteristics; assess the predictive value of digital mobility measures for adverse outcomes; and model risk factors for the onset and progression of mobility decline in older Canadians. To allow for trajectory analyses and to account for potential seasonal changes in activity patterns, we incorporated frequent repeated follow-ups (every 4 months).

Based on our original objectives, our target sample size was 1,500 participants, grounded in both feasibility and predictive validity considerations. Prior studies examining the accuracy of mobility measures for predicting health outcomes have reported area under the curve (AUC) values ranging from 0.6 to 0.8.^30,31^ To ensure adequate power (80%) to detect a significant deviation from the null hypothesis (AUC = 0.5; two-sided test at p < 0.05), we based our calculations on the most conservative expected AUC of 0.6. Based on conservative expected incidence rates for key outcomes: at least 1 fall (20% over 1 year), visit to the emergency room (12% over 1 year), overnight hospital stays (8% over 2 years), and progression to major mobility disability (10% over 2 years), a sample of approximately 740 participants would be needed to detect progression to major mobility limitation over two years, and around 634 participants to detect emergency room visits over the same period. Accounting for an anticipated dropout rate of 20% and the potential for missing device-based data of up to 20%, the required sample increases to approximately 1,156 for progression to major mobility disability and 991 for ER visits. This sample size is also sufficient for detecting small and clinically relevant differences between the three mobility level groups based on the PCML for step count (400-600 steps) and for trips per week (4-6 trips). Our flowchart (Figure 1) updated on September 23, 2025, highlights that we completed our 12-month follow-up with 1,378 active participants.

**Figure 1.**
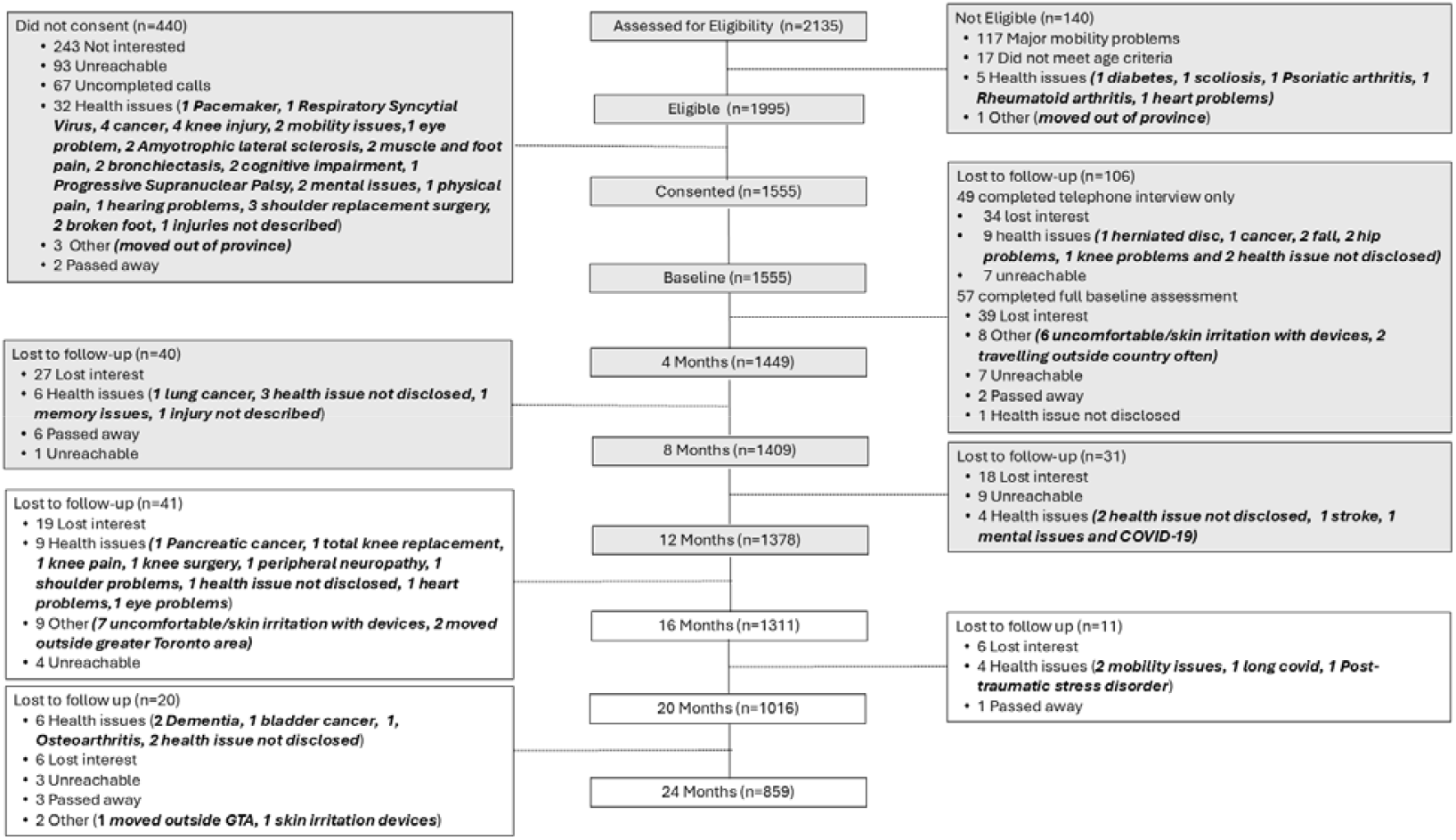
Flow of study participants and timeline of data collection in the MacM3 study, from recruitment through September 2025. Grey areas indicate completed follow-up periods.

### Study Procedures

Participants in MacM3 are asked to wear a consumer grade smartwatch with Global Navigation Satellite System (GNSS) and activity monitoring capabilities, the TicWatch Pro 3 Ultra GPS^27^ on the wrist for 10 days at baseline and at 4-month intervals for a total of seven time points of data collection over 24 months. At the time of consent, participants were also asked if they would be willing to wear a second (optional) activity monitoring device, the ActiGraph wGT3X-BT^27,32^ or the Axivity X3,^33^ on the thigh concurrently with the wrist-worn device. At baseline, 12 months, and 24 months, participants complete a more in-depth series of questionnaires about their health and mobility through a combination of telephone interviews and online surveys as well as an in-person physical assessment. In between the yearly assessments, devices are mailed to participants’ homes every 4 months and then returned to the study site via pre-paid shipping. To monitor health outcomes (e.g., new diagnoses, healthcare utilization, injuries, falls), we use quarterly phone calls and monthly mail-in fall diaries.^34^ Figure 2 provides an overview of the MacM3 study design.

**Figure 2.**
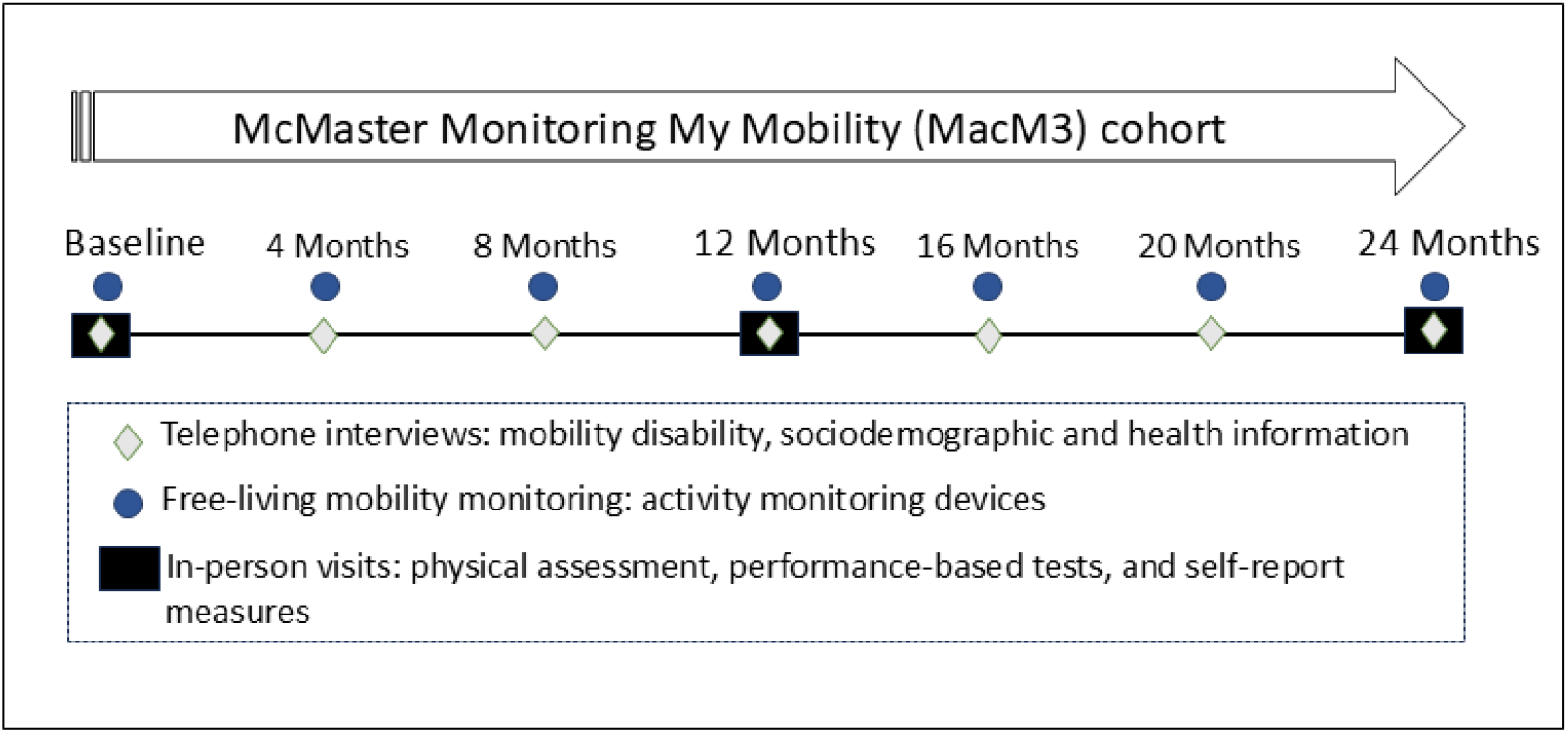
MacM3 study overview.

### Study measures

Our data collection encompasses all three “tenses” of mobility/function in line with the model first proposed by Glass^35^ and outlined more recently by members of our team based on the World Report on Ageing and Health^36^: enacted (i.e., actual mobility in the community measured via wearables), hypothetical (i.e., perceived ability measured via self-reported questionnaires), and experimental (i.e., locomotor capacity for mobility measured via in-person physical assessments). Self-reported measures of mobility provide information on what patients “can do”; performance-based testing provides information on what patients “could do”; and importantly, the sensor data on enacted/actual mobility provide information on what patients actually “do” in the real-world.^36^

### Mobility Measures

#### Enacted mobility

The MacM3 study employs a multi-sensor, multi-nodal approach for studying enacted and free-living mobility, integrating time-synchronized accelerometry with GPS in a wrist-worn device alongside a second thigh-worn accelerometer.^27^ Device selections and wear locations for the current cohort were made after extensive lab-based testing, consultation with our older adult Stakeholder Advisory Committee, and pilot testing to determine their reliability and technical validity.^27^

Participants are asked to wear a TicWatch Pro 3 Ultra GPS on their non-dominant wrist and either the ActiGraph GT3X-BT or an Axivity AX3 on the thigh for 10 consecutive days during waking hours at baseline and every 4 months throughout the study. The **wrist-worn device** (TicWatch by Mobvoi) is a smartwatch that uses the Wear OS operating system by Google™ and has a built-in tri-axial accelerometer, gyroscope, heart rate sensor, and multi-GNSS (GPS, GLONASS, Galileo, QZSS, and Beidou) for logging position coordinates and time. As the watch was intended to be used in our study as a stand-alone data logging device without a cellphone, GNSS data are collected at different intervals depending on the predicted activity state of the device using Google’s Activity Recognition API. To improve battery life, we configured the device to collect location coordinates every 5 seconds when the watch is in an active motion state (e.g., walking, in a vehicle) with good satellite reception, and less frequently when the device is still or there is poor satellite reception.^27^ We validated the TicWatch against a gold-standard stand-alone GPS device (Qstarz GPS) and validated its accelerometer output against the thigh-worn ActiGraph device. In this validation study, we found that the TicWatch models were valid devices for capturing both GPS and raw accelerometer data, supporting their use as reliable tools for assessing real-life mobility in older adults.^27^

The second device is **a thigh-worn accelerometer** (either the ActiGraph wGT3X-BT or Axivity AX3) secured to the upper thigh using an adhesive patch (Hypafix). The ActiGraph wGT3X-BT^37,38^ (ActiGraph, Pensacola, FL) and Axivity AX3^33^ (Axivity Ltd, United Kingdom) are commonly used in epidemiological research for measuring physical activity ^38–40^ and body posture, with thigh placement considered the gold standard for accurately distinguishing between sitting, standing, and walking.^27,41–44^ Both devices have built-in triaxial accelerometers for recording high-resolution raw acceleration data and long-lasting batteries that eliminate the need for recharging during the data collection period. In our validation study, the ActiGraph demonstrated high accuracy in detecting body posture. Full details of this validation study have been published elsewhere.^27^

A custom data collection application, Ivy, was designed by our team exclusively for the TicWatch to optimize data quality and battery life by collecting data from onboard sensors, including the accelerometer, gyroscope, geolocation, and heart rate.^27^ A companion app, Clover, supports Ivy on Windows by automating the setup of both the TicWatch and the Axivity device, streamlining data downloads and device preparation. The ActiGraph device, on the other hand, is initialized using the ActiLife proprietary software. Additionally, Clover integrates information on the date, time, and completion status of data downloads with our REDCap data collection system for all devices, including the ActiGraph.

The primary digitally-derived mobility measures of interest for MacM3 can be classified into five core domains: *activity* (amount of time spent in sedentary, light, moderate and vigorous physical activity),^45,46^ *volume-related features of walking* (steps/day, walking bout lengths and duration), *spatiotemporal features of walking* (gait cadence, variability, velocity, and stride width during long bouts),^47^ *postural* (time spent lying down, sitting, standing, and frequency of transitions),^42,48^ and *geospatial features* (trip frequency, trip duration, trip mode, maximum distance from home, and minimum convex hull/life space area).^49,50^ The derivation of these digital mobility measures has been described in previous technical validation papers by our team and others.^36,51–53^ Notably, with integrated GNSS and accelerometry data, we are able to uniquely distinguish and characterize indoor and outdoor mobility (bouts, trips) and patterns of mobility outside of the home.

All measures are processed using the NiMBaLWear^51^ pipeline which is a modular, open-source wearable sensor analytic framework designed to accommodate a wide range of sensors, signal types, and body wear locations. NiMBalWear was developed to address the limitations of device-specific processing by providing a standardized, flexible approach for analyzing raw accelerometry data across different platforms. This framework unifies the measurement and analysis of multiple health-related domains—such as mobility, posture, and physical activity—by optimizing data extraction from individual sensors and enabling data fusion across multiple sensor types and locations when applicable. By focusing on raw signal processing, NiMBaLWear ensures device-agnostic harmonization during the feature extraction stage, allowing for consistent, comparable metrics regardless of brand or configuration. This is particularly important for longitudinal studies or multi-site research using different devices, as it supports reproducible and scalable analysis without reliance on proprietary algorithms or cut-points. Currently, the NiMBaLWear platform also includes a Mobility module focused on IMU-based outcomes, which is being expanded to incorporate GNSS+ analytics.

##### NiMBaLWear-based processing of thigh- and wrist-worn accelerometry data

Step counts and walking bouts are derived from the thigh-worn accelerometer. NiMBaLWear applies wavelet-based time-frequency analysis to the triaxial acceleration signal to detect unilateral steps.^54,55^ Step-related events are identified based on the frequency content of the acceleration profile, capturing both the swing and stance phases of walking. Foot contact is used to define the timing of each step following a detected swing phase. Consecutive steps are then grouped into walking bouts according to predefined timing criteria (e.g., a minimum number of steps within a maximum interval). This method offers greater precision than traditional threshold-based techniques and has demonstrated improved accuracy in detecting steps and walking patterns, particularly in older adults. The step count data reported in this manuscript aligns with findings from other large cohort studies^56,57^ that applied machine learning methods to estimate step count from wrist-worn devices and observed similar trends—namely, a decline in step counts with advancing age and increased mobility limitations.

Postural behaviour is derived from the same thigh-worn triaxial accelerometer using algorithms based on the orientation of the device relative to gravity. Specifically, posture was classified as either standing or sitting/lying based on the acceleration due to gravity recorded along each axis of the accelerometer, assuming consistent device placement on the thigh. Postural transitions between standing and sitting/lying were detected by identifying periods of sustained change in thigh orientation that met predefined criteria. Once a transition was detected, all subsequent time points were classified according to the last identified posture. This method allows us to determine body posture continuously throughout the wear period using a single device, with classification based solely on orientation and transition patterns.

Activity intensity, including sedentary behavior (SED), light physical activity (LPA), and moderate-to-vigorous physical activity (MVPA), is derived from the wrist device. We applied epoch-independent cut-points validated by Fraysse et al.,^58^ which classify activity based on raw acceleration (in mg). Specifically, thresholds of 42.5 mg (non-dominant wrist) and 62.5 mg (dominant wrist) were used for light activity, while 98.0 mg (non-dominant) and 92.5 mg (dominant) defined moderate activity. These cut-points have been applied in other aging cohorts and support comparability across studies examining accelerometry-derived activity patterns.

#### Hypothetical mobility

Perceived mobility is assessed through various self-report instruments developed to capture different mobility domains among older adults.^59,60^ The Late-Life Function and Disability Instrument (LLFDI)^61,62^ evaluates both physical function mobility and participation in valued life roles, encompassing tasks from basic physical actions to community activities. The Mobility Assessment Tool – short form (MAT-sf)^63^ is a 10-item computer-administered measure of perceived mobility using video technology. The Life-Space Assessment (LSA)^64^ measures life-space mobility – the distance, frequency and independence perceived by individuals as they move through their communities. Transportation mobility is evaluated through questions from the Canadian Longitudinal Study on Aging (CLSA)^65^ Transportation, Mobility, Migration (TRA)^66^ module, while driving behavior is assessed using the Driving Questionnaire, Driving Comfort Scale (DCS)^67,68^ as well as selected questions from the Perceived Driving Abilities (PDA) Scale.^67^

#### Experimental mobility

Participants undergo in-person physical assessments at either the Hamilton or Toronto location at baseline, 12-months, and 24-months follow-up. These assessments include commonly used and well-validated performance-based tests of mobility and physical function, namely the Timed Up and Go (TUG)^69^ usual and dual-task tests, the repeated sit-to-stand test (5 x STS), tests of gait speed^70^ (both usual and fast pace) and single-leg balance^71^. The 400-meter walk test^72,73^ evaluates aerobic capacity, while muscle power is estimated with the stair-climbing power test.^74,75^ Other physical measures collected at the in-person assessment include the Clinical Frailty Scale (CFS),^76^ Visual Acuity (VA) test,^77^ spirometry^78^ for pulmonary function evaluation, and handgrip strength^79^ assessment. Participants are permitted to use their mobility aids during the TUG, gait speed, stair-climbing, and 400-meter walk tests. Use of mobility aids was documented and recorded in the study database for future analyses.

#### Demographic and health measures

As part of the yearly in-person assessments, participants also complete a series of questionnaires related to their basic health and the possible physical, psychosocial, cognitive, financial, and environmental contributors to mobility. This encompasses demographics, such as socioeconomic status and lifestyle factors, as well as health-related behaviors. We assess physical activity through the Physical Activity Scale for the Elderly (PASE),^80^ nutrition risk using the Seniors in the Community: Risk Evaluation for Eating and Nutrition (SCREEN-8),^81^ pain using the Brief Pain Inventory-Short Form^82^, and health status with the EuroQol-5D-5L (EQ-5D-5L).^83^ Additionally, cognitive^84^ and hearing^85^ function, social support,^86–88^ and mental health^89,90^ are assessed through validated tools. These measures are captured using a combination of telephone interviews and online surveys at baseline, 12-month, and 24-month follow-ups.

To monitor health and mobility outcomes longitudinally, participants are contacted at 4-month intervals from baseline until the conclusion of the study. During these calls, the PCML^91^ is re-administered to track changes over time and to identify the development of mobility disability. Participants also rate their perceived changes in mobility using the Global Rating of Change (GRC)^92^ scale and answer questions about healthcare utilization, including hospitalizations, emergency room visits, primary care visits, rehabilitation service use, motor vehicle collisions, new diagnoses, living situation updates, and general and mental health status.

Falls are measured prospectively using monthly fall diary calendars, which are provided to participants along with pre-addressed and pre-stamped postcards. This method of data collection for falls is regarded as the gold standard by international expert consensus.^34^ As part of the data quality checks, the research team calls participants to rectify missing data and confirm reported falls, ensuring they fit the definition of a fall^34,93^ and documenting circumstances. Participants who report falls are requested to complete a standardized case report form via telephone to collect detailed information, with fall-related injuries categorized as serious, moderate, or minor based on symptoms and healthcare use. Participants are encouraged to report injurious falls promptly for accurate documentation. Table 1 provides an overview of the measures and corresponding assessment time points.

### Patient and Public Involvement

Older adult stakeholders (n=7) were actively engaged throughout the development of the MacM3 cohort study. They provided valuable feedback on various aspects of the project, including the study concept (interest and perceived value of mobility monitoring) and data collection methods (i.e., selection of the wearable device and wear location). While older adult stakeholders were not directly involved in defining the research questions or selecting outcome measures, their feedback informed the perceived value, feasibility, and burden of study procedures.

We plan to involve both stakeholder and community advisory groups in the interpretation and dissemination of study findings, including selecting relevant information and appropriate formats for sharing results with participants and wider communities. We will continue to consult with our adult partners regarding the interpretation of study findings, and strategies for translating research outcomes into practical applications. This ongoing engagement helps ensure the research remains relevant to the needs and concerns of the community it seeks to benefit. To maintain this strong connection, the project plans to meet with stakeholders at least once a year, offering a forum for continued dialogue and input.

As part of our partnership with the Dixon Hall Centre in Toronto, we also consulted extensively with older adults and social service providers in the community to ensure our data collection methods were as inclusive as possible. A community advisory group was assembled (11 members) who continue to provide input on all stages of our research at the Centre which has resulted in key modifications to our protocol to facilitate participant recruitment and retention including: options for retuning the device in person, Mandarin speaking assessors, drop-in hours for trouble shooting, gift cards for returning devices on time, option to complete online survey in-person, and distributing flyers that outlined our commitment to addressing the underrepresentation of diverse older adults in research—while also emphasizing safety and security. This comprehensive approach to stakeholder involvement ensures that the MacM3 cohort reflects a wide range of experiences and perspectives, strengthening the overall impact and generalizability of the research.

## Results

### Participants characteristics

We aimed to enroll a total of 1500 participants, and as of May 2024, recruitment was completed with 1210 participants enrolled at the Hamilton site and 345 at the Dixon Hall site, for a total of 1555 participants. Figure 3 displays the geographic distribution of participant addresses, demonstrating a concentration of recruitment in Hamilton and Toronto, Ontario, aligning with the study’s targeted catchment area. The inset highlights the dense and well-distributed recruitment pattern within Hamilton, indicating strong geographic coverage across the city. Based on Statistics Canada’s Population Centre and Rural Area Classification, most participants (n = 1,416) reside in large population centers. A smaller number live in medium (n = 5) and small (n = 57) population centers, and 76 participants were recruited from rural areas. One participant’s address was missing and could not be classified.

**Figure 3.**
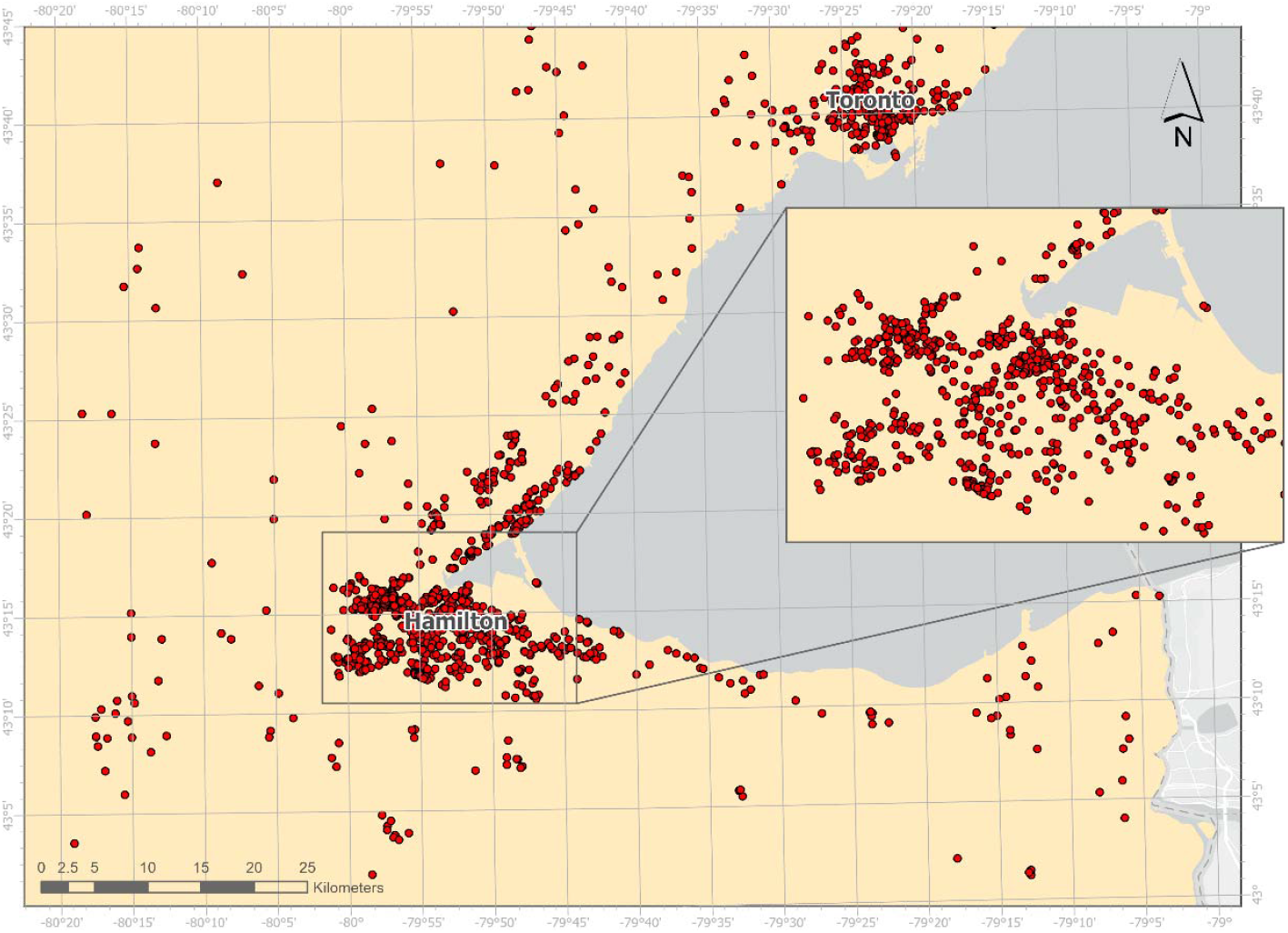
Geographic distribution of MacM3 participants.

Figure 1 illustrates the flow of study participants from recruitment through each follow-up period, highlighting participant retention and attrition trends over time. A total of 2,135 individuals expressed interest in the MacM3 study. Of these, 140 were deemed ineligible— primarily due to major mobility limitations (n=117), based on our PCML screening survey. Additionally, 440 individuals declined to participate. As a result, 1,555 participants consented to the MacM3 study. Baseline assessments were completed for all enrolled participants, along with 4-, 8-, and 12-month follow-ups, with 1,378 participants completing the 12-month assessment. The study continues to demonstrate good retention, with an overall attrition rate of just 16.0%. The highest dropout rate occurred at the beginning of the study, between baseline and 4 months, where many participants (n=57, 3.7%) dropped out after completing all components of the baseline assessment. The most common reasons for dropout included participants losing interest, becoming unreachable, device discomfort or experiencing health-related challenges. Mortality has been reported in 12 participants (0.7%) during follow-up to date. As of the latest follow-up, 859 participants have successfully completed their24-month follow-up; data collection is still ongoing. The study’s overall high retention rate underscores the feasibility of long-term data collection using wearable devices and highlights the effectiveness of our engagement strategies in sustaining participant involvement

Table 2 presents a comprehensive overview of the sociodemographic, lifestyle, and health characteristics of the MacM3 participants at baseline. Among the total cohort (n = 1555), 68.4% were female, and the mean age at baseline was 73.9 years (SD = 5.5). A majority of participants (62.4%) were married or living with a partner, and 75.3% had attained a post-secondary diploma. Additionally, 59.8% reported never having smoked, and 83.3% were regular drinkers. In terms of health-related measures, 62.9% reported having three or more comorbidities, with cataracts, osteoarthritis and hypertension being the most reported conditions. Furthermore, 81.2% were taking prescribed medication(s), and 44.2% and 41.6% rated their general health and mental health, respectively, as ‘very good.’ Further details of the cohort are provided in Supplement 1.

**Table 2.** Selected baseline characteristics of the MacM3 participants.

|  |  | Total | Hamilton site |  | Dixon Hall site |  |
| --- | --- | --- | --- | --- | --- | --- |
|  |  |  | Males<br>*n* (%) | Females<br>*n* (%) | Males<br>*n* (%) | Females<br>*n* (%) |
| Sample size, *n* (%) |  | 1555 | 408 (33.7) | 802 (66.3) | 83 (24.0) | 262 (76.0) |
| Age (years), mean (SD) |  | 73.9 (5.5) | 74.4 (5.7) | 73.5 (5.4) | 74.8 (5.8) | 73.8 (5.5) |
| BMI (kg/m<sup>2</sup>), mean (SD) |  | 26.3 (4.8) | 27.0 (4.2) | 26.4 (4.9) | 25.8 (5.9) | 25.2 (5.0) |
| Country of Birth, *n* (%) | Born in Canada | 1,079 (69.4) | 296 (72.6) | 621 (77.4) | 35 (42.2) | 127 (48.5) |
| Ethnicity (*participants could select multiple ethnicities*) | White/Caucasian | 1369 (88.0) | 388 (95.1) | 770 (96.0) | 48 (57.8) | 163 (62.2) |
|  | Chinese | 108 (7.0) | 1 (0.2) | 4 (0.5) | 32 (38.6) | 71 (27.1) |
|  | Other (Black, Japanese, South Asian, Latin American, North American Indian, Metis, Arab, Korean, other) | 88 (5.7) | 25 (6.1) | 31 (3.9) | 2 (2.4) | 30 (11.4) |
| Marital Status, *n* (%) | Married/living with a partner | 970 (62.4) | 337 (82.6) | 457 (57.0) | 67 (80.8) | 109 (41.6) |
|  | Widowed | 229 (14.7) | 22 (5.4) | 158 (19.7) | 4 (4.8) | 45 (17.2) |
|  | Divorced | 174 (11.2) | 15 (3.7) | 110 (13.8) | 8 (9.6) | 41 (15.6) |
|  | Separated | 36 (2.3) | 9 (2.2) | 16 (2.0) | 0 (0.0) | 11 (4.2) |
|  | Single | 145 (9.3) | 25 (6.1) | 60 (7.5) | 4 (4.8) | 56 (21.4) |
| Education, *n* (%) | Secondary school incomplete | 54 (3.4) | 12 (2.9) | 26 (3.3) | 2 (2.4) | 14 (5.4) |
|  | Secondary school completed | 158 (10.2) | 53 (13.0) | 74 (9.2) | 6 (7.2) | 25 (9.5) |
|  | Some post-secondary education | 172 (11.1) | 44 (10.8) | 103 (12.8) | 7 (8.5) | 18 (6.9) |
|  | Post-secondary diploma | 1,171 (75.3) | 299 (73.3) | 599 (74.7) | 68 (81.9) | 205 (78.2) |
| *Work status, *n* (%) |  |  |  |  |  |  |
| (*participants could select multiple work status*) | Retired | 1084 (69.7) | 285 (69.6) | 542 (67.6) | 60 (72.3) | 197 (75.2) |
|  | Working (part time or full time) | 125 (8.0) | 37 (9.1) | 57 (7.1) | 11 (13.5) | 20 (7.6) |
|  | Volunteer | 75 (4.8) | 7 (1.7) | 27 (3.4) | 11 (13.5) | 30 (11.5) |
|  | Other | 37 (2.4) | 9 (2.2) | 16 (2.0) | 2 (2.4) | 14 (5.3) |
| Household income, *n* (%) | < \$20,000 | 86 (5.5) | 3 (0.7) | 5 (0.6) | 21 (25.3) | 57 (21.8) |
| | \$20,000-49,999 | 331 (21.3) | 57 (14.0) | 189 (23.6) | 15 (18.1) | 70 (26.7) |
| | \$50,000-\$99,999 | 654 (42.1) | 184 (45.1) | 370 (46.1) | 20 (24.1) | 80 (30.5) |
| | ≥ \$100,000 | 399 (25.7) | 147 (36.0) | 178 (22.2) | 26 (31.3) | 48 (18.3) |
|  | Don't know/Prefer not to answer | 85 (5.5) | 17 (4.2) | 60 (7.5) | 1 (1.2) | 7 (2.7) |
| Smoking status, *n* (%) | Current smoker | 31 (2.0) | 11 (2.7) | 16 (2.0) | 1 (1.2) | 3 (1.1) |
|  | Former smoker | 594 (38.2) | 195 (47.8) | 298 (37.2) | 36 (43.4) | 65 (24.8) |
|  | Never smoked | 930 (59.8) | 202 (49.5) | 488 (60.8) | 46 (55.4) | 194 (74.1) |
| Alcohol consumption past year, *n* (%) | Regular drinker (> once a month) | 1,041 (83.3) | 315 (88.0) | 552 (81.5) | 42 (87.5) | 132 (79.0) |
|  | Occasional drinker (< once a month) | 206 (16.5) | 42 (11.7) | 123 (18.2) | 6 (12.5) | 35 (21.0) |
|  | Non-drinker | 3 (0.2) | 1 (0.3) | 2 (0.3) | 0 (0.0) | 0 (0.0) |
| Comorbidities, *n* (%) | None | 54 (3.5) | 21 (5.1) | 21 (2.6) | 2 (2.4) | 10 (3.8) |
|  | < 3 | 522 (33.6) | 126 (30.9) | 270 (33.7) | 30 (36.1) | 96 (36.7) |
|  | ≥ 3 | 979 (62.9) | 261 (64.0) | 511 (63.7) | 51 (61.5) | 156 (59.5) |
| Prescribed medication, *n* (%) | Yes | 1,261 (81.2) | 355 (87.5) | 652 (81.4) | 71 (85.5) | 183 (69.8) |
|  | No | 291 (18.8) | 51 (12.5) | 149 (18.6) | 12 (14.5) | 79 (30.2) |

BMI: body mass index; SD: standard deviation; \*Data on working status was added later to the protocol, and data collection for this variable is still ongoing.

More than two-thirds of the participants were born in Canada (69.4%). Most identified as White/Caucasian (88.0%), with smaller proportions identifying as Chinese (7.0%) or another ethnic background such as Black, Japanese, South Asian, Latin American, North American Indigenous, Métis, Arab, or Korean (5.7%). Ethnic diversity was more pronounced at the Dixon Hall site, where less than half of the participants were born in Canada. In terms of cultural background, participants frequently reported English (40.0%), Scottish (25.1%), Irish (24.1%), and Canadian (14.5%) heritage, with others identifying German, Chinese, French, Italian, Polish, Dutch, or other cultural backgrounds. Cultural diversity was also more evident at the Dixon Hall site, where 38.6% of male participants and 27.1% of females reported Chinese cultural heritage, and a wide range of other cultural identities were selected, including Ukrainian, Métis, and North American Indigenous backgrounds (Supplement 1).

### Findings to date

Table 3 describes the self-reported mobility and performance-based test results at baseline. Most participants (93.8%) did not use mobility aids, and 37.6% reported experiencing a fall in the previous year. According to the PCML classification, 56.7% of participants had no mobility limitations, while 15.9% had preclinical mobility limitations, and 27.4% had minor mobility limitations at baseline.

**Table 3.** Baseline self-reported mobility, falls and performance-based tests results.

|  |  | Total Sample<br>*n* (%) | Males<br>*n* (%) | Females<br>*n* (%) |
| --- | --- | --- | --- | --- |
| Self-reported mobility, *n* |  | 1555 | 491 | 1064 |
| Preclinical Mobility Disability Scale | No mobility limitation | 881 (56.7) | 298 (60.7) | 583 (54.8) |
|  | Preclinical mobility limitation | 248 (15.9) | 67 (13.6) | 181 (17.0) |
|  | Minor mobility limitation | 426 (27.4) | 126 (25.7) | 300 (28.2) |
| Mobility aids, *n* |  | 1553 | 490 | 1063 |
|  | Yes | 96 (6.2) | 30 (6.1) | 66 (6.2) |
|  | No | 1457 (93.8) | 460 (93.9) | 997 (93.8) |
| Falls in the past year, *n* |  | 1555 | 491 | 1064 |
|  | Yes | 584 (37.6) | 169 (34.4) | 415 (39.0) |
|  | No | 971 (62.4) | 322 (65.6) | 649 (61.0) |
| Performance-based tests |  |  |  |  |
|  | <sup>a</sup>Grip Strength (Kg), *n* | 1504 | 479 | 1025 |
|  | mean ± SD | 27.3 ± 9.4 | 37.2 ± 8.3 | 22.7 ± 5.5 |
|  | Gait speed (m/s), *n* | 1506 | 480 | 1026 |
|  | mean ± SD | 1.23 ± 0.2 | 1.24 ± 0.2 | 1.22 ± 0.2 |
|  | Timed up and go (s), *n* | 1505 | 479 | 1026 |
|  | mean ± SD | 9.4 ± 2.5 | 9.5 ± 2.5 | 9.4 ± 2.6 |
|  | 5 × sit-to-stand (s), *n* | 1472 | 466 | 1006 |
|  | mean ± SD | 13.0 ± 3.8 | 12.9 ± 3.5 | 13.0 ± 4.0 |
|  | <sup>a</sup>Single leg stance (s), *n* | 1467 | 463 | 1004 |
|  | mean ± SD | 29.6 ± 22.7 | 26.4 ± 22.1 | 31.1 ± 22.8 |
|  | <sup>b</sup>400 meters walk test (min), *n* | 1109 | 381 | 728 |
|  | mean ± SD | 4.9 ± 0.9 | 4.8 ± 1.0 | 4.9 ± 0.9 |
|  | <sup>b</sup>Stair climbing Power (W), *n* | 1140 | 389 | 751 |
|  | mean ± SD | 258.1 ± 84.0 | 313.3 ± 89.9 | 229.5 ± 64.2 |

<sup>a</sup>best of left and right test; <sup>b</sup>The 400 meters walk test and the stair climbing power test are not conducted at DH site.; W = watts

For the performance-based tests, the mean of the best right- or left-hand grip strength in the overall cohort was 27.3 kg (SD = 9.4). The mean gait speed was 1.23 m/s (SD = 0.02), the mean Timed Up and Go test time was 9.4 seconds (SD = 2.5), and the mean 5x sit-to-stand test time was 13.0 seconds (SD = 3.8). Results are also stratified by sex. Males generally demonstrated higher grip strength and stair climbing power, while other functional measures were relatively consistent across groups. Overall, these results reflect a relatively high-functioning sample of older adults at baseline, which is consistent with our recruitment strategy.^94^

#### Preliminary results - wearables data

At baseline, data from 1412 participants were processed. Wear time in our study was defined as at least 10 hours per day on a minimum of 7 days. Based on this criterion, 62 participants had no valid wrist wear data, and 49 wore the wrist device for fewer than 7 valid days. A total of 1,301 participants had valid wrist-worn device data, with an average wear time of 13.6 hours per day (range: 7.2 to 18.3 hours) over 10.2 days. For the thigh-worn device, 309 participants declined to wear it at baseline, and 95 did not meet the wear-time criteria. As a result, data from 1,008 participants are presented. On average, participants wore the thigh device for 13.6 hours per day (range: 6.1 to 22.5 hours) over approximately 10.6 days.

Table 4 presents the mean and median daily step counts by quartile. Step counts ranged from 5,073 steps in the lowest quartile to 12,303 steps in the highest. Participants with higher activity levels (Q3: 72.9 years; Q4: 72.7 years) were slightly younger than those with lower activity (Q1: 74.6 years; Q2: 74.7 years). Mobility status also varied with activity level: 73.8% of those in the highest quartile reported no mobility limitations, versus only 36.9% in the lowest quartile. Wear-time compliance was consistent across all quartiles, with a median of 10 compliant days and average daily wear time ranging from 13.8 to 14.6 hours. These data are consistent with findings from other large cohort studies showing a decline in step counts with advancing age and increasing mobility limitations.^95,96^

**Table 4.** Baseline characteristics of participants by quartiles of mean steps per day derived from the thigh devices (n=1008).

| Characteristics | Total Sample | Quartiles of mean steps per day, mean (SD) |  |  |  |
| --- | --- | --- | --- | --- | --- |
|  |  | Quartile 1 (Lowest) | Quartile 2 | Quartile 3 | Quartile 4 (Highest) |
| Total participants, n | 1008 | 252 | 252 | 252 | 252 |
| Age (years), mean (SD) | 73.7 (5.4) | 74.6 (5.5) | 74.7 (5.5) | 72.9 (5.2) | 72.7 (4.9) |
| Mobility score, n (%) |  |  |  |  |  |
| No mob limitation | 582 (57.7) | 93 (36.9) | 149 (59.1) | 154 (61.1) | 186 (73.8) |
| Preclinical | 166 (16.5) | 49 (19.4) | 41 (16.3) | 47 (18.7) | 29 (11.5) |
| Minor mob limitation | 260 (25.8) | 110 (43.7) | 62 (24.6) | 51 (20.2) | 37 (14.7) |
| No. of Days with wear ($\geq 7$ days) | | | | | |
| Mean (SD) | 9.9 (1.2) | 9.7 (1.2) | 9.8 (1.0) | 10.0 (1.5) | 10.0 (1.0) |
| median | 10 | 10 | 10 | 10 | 10 |
| Hours of wear on compliant days ($\geq 10$h) | | | | | |
| Mean (SD) | 14.1 (1.4) | 13.8 (1.4) | 14.1 (1.3) | 14.1 (1.3) | 14.6 (1.2) |
| Median | 14.1 | 13.7 | 14.0 | 14.1 | 14.6 |
| Steps/day |  |  |  |  |  |
| Mean (SD) | 8437 (2943) | 5073 (962) | 7306 (490) | 9065 (560) | 12303 (2283) |
| Median | 8175 | 5278 | 7318 | 8971 | 11694 |

Trip data from 1,422 participants were processed using the algorithms proposed by Fillekes et al.^27,49^, as described in Table 5. Applying the study’s compliance criteria of ≥7 days of wrist wear with ≥10 hours per day, data from 1,323 participants are presented. On average participants wore the wrist device for 12.1 (SD = 1.3) hours. In terms of mobility outcomes, participants recorded an average of 2.0 active trips per day (SD = 1.3) and 1.8 passive trips per day (SD = 1.2). Active and passive trips ranged from fewer than one trip in the lowest quartile to over three trips in the highest quartiles. Participants in the highest quartile were, on average, younger and had fewer mobility limitations.

**Table 5.**
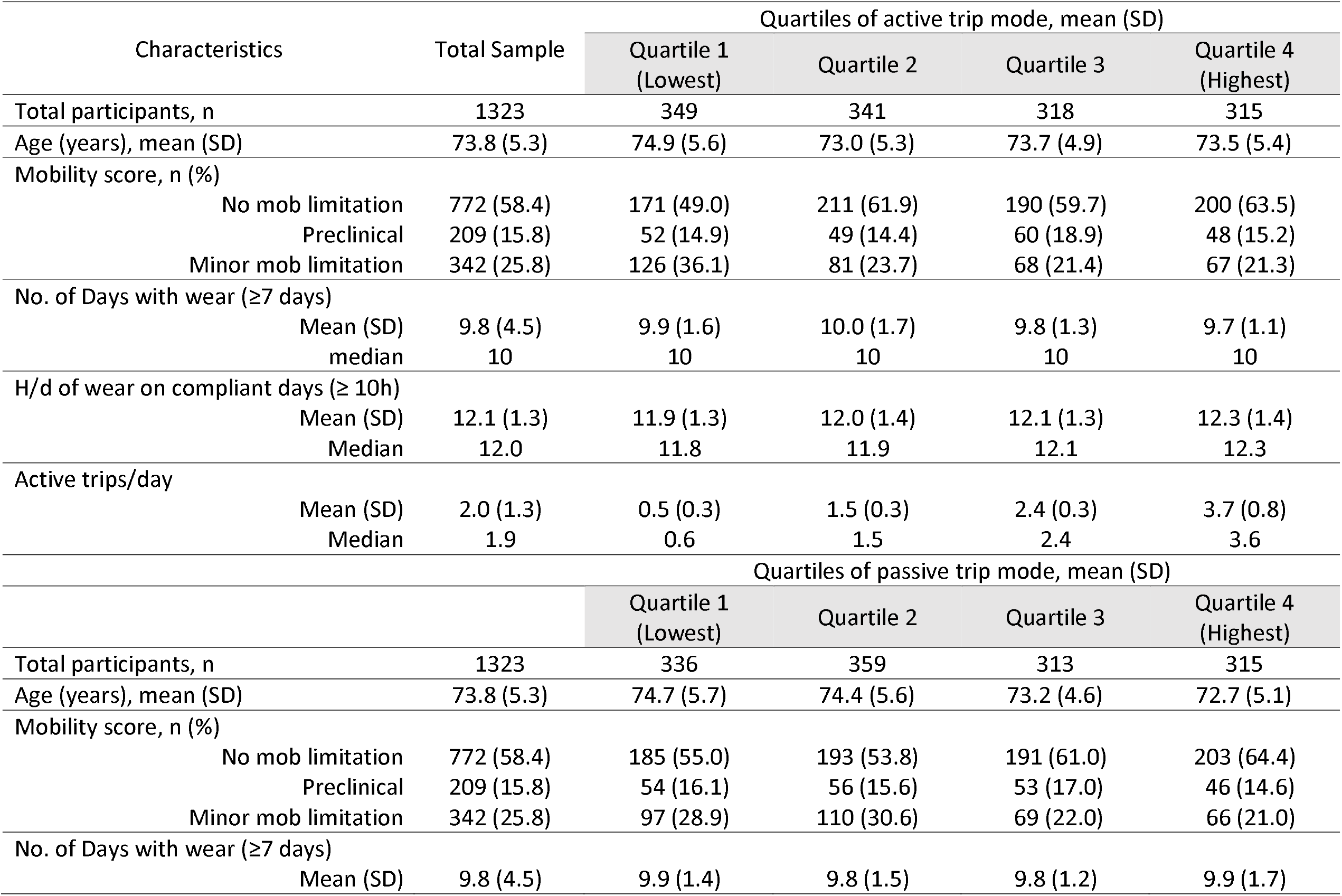
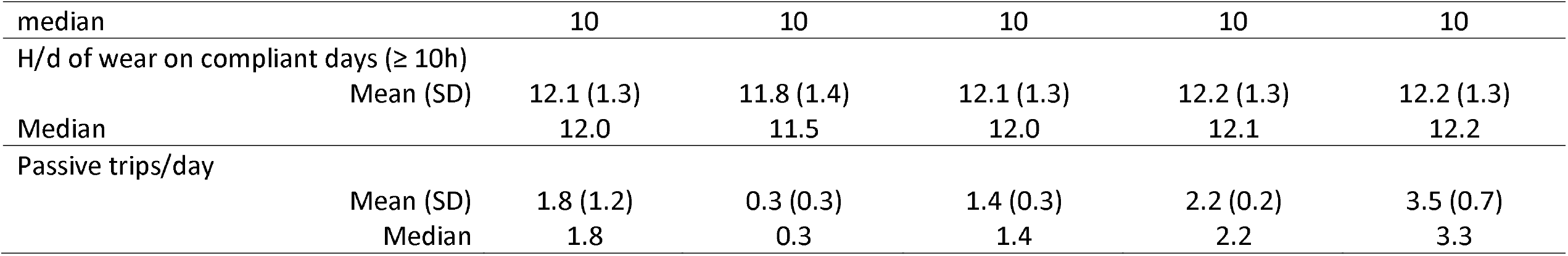
Baseline characteristics of trip mode derived from the wrist device (n=1323).

| Characteristics | Total Sample | Quartiles of active trip mode, mean (SD) |  |  |  |
| --- | --- | --- | --- | --- | --- |
|  |  | Quartile 1<br>(Lowest) | Quartile 2 | Quartile 3 | Quartile 4<br>(Highest) |
| Total participants, n | 1323 | 349 | 341 | 318 | 315 |
| Age (years), mean (SD) | 73.8 (5.3) | 74.9 (5.6) | 73.0 (5.3) | 73.7 (4.9) | 73.5 (5.4) |
| Mobility score, n (%) |  |  |  |  |  |
| No mob limitation | 772 (58.4) | 171 (49.0) | 211 (61.9) | 190 (59.7) | 200 (63.5) |
| Preclinical | 209 (15.8) | 52 (14.9) | 49 (14.4) | 60 (18.9) | 48 (15.2) |
| Minor mob limitation | 342 (25.8) | 126 (36.1) | 81 (23.7) | 68 (21.4) | 67 (21.3) |
| No. of Days with wear (≥7 days) |  |  |  |  |  |
| Mean (SD) | 9.8 (4.5) | 9.9 (1.6) | 10.0 (1.7) | 9.8 (1.3) | 9.7 (1.1) |
| median | 10 | 10 | 10 | 10 | 10 |
| H/d of wear on compliant days (≥ 10h) |  |  |  |  |  |
| Mean (SD) | 12.1 (1.3) | 11.9 (1.3) | 12.0 (1.4) | 12.1 (1.3) | 12.3 (1.4) |
| Median | 12.0 | 11.8 | 11.9 | 12.1 | 12.3 |
| Active trips/day |  |  |  |  |  |
| Mean (SD) | 2.0 (1.3) | 0.5 (0.3) | 1.5 (0.3) | 2.4 (0.3) | 3.7 (0.8) |
| Median | 1.9 | 0.6 | 1.5 | 2.4 | 3.6 |
|  |  | Quartiles of passive trip mode, mean (SD) |  |  |  |
|  |  | Quartile 1<br>(Lowest) | Quartile 2 | Quartile 3 | Quartile 4<br>(Highest) |
| Total participants, n | 1323 | 336 | 359 | 313 | 315 |
| Age (years), mean (SD) | 73.8 (5.3) | 74.7 (5.7) | 74.4 (5.6) | 73.2 (4.6) | 72.7 (5.1) |
| Mobility score, n (%) |  |  |  |  |  |
| No mob limitation | 772 (58.4) | 185 (55.0) | 193 (53.8) | 191 (61.0) | 203 (64.4) |
| Preclinical | 209 (15.8) | 54 (16.1) | 56 (15.6) | 53 (17.0) | 46 (14.6) |
| Minor mob limitation | 342 (25.8) | 97 (28.9) | 110 (30.6) | 69 (22.0) | 66 (21.0) |
| No. of Days with wear (≥7 days) |  |  |  |  |  |
| Mean (SD) | 9.8 (4.5) | 9.9 (1.4) | 9.8 (1.5) | 9.8 (1.2) | 9.9 (1.7) |

|  |  |  |  |  |  |  |
| --- | --- | --- | --- | --- | --- | --- |
| median |  | 10 | 10 | 10 | 10 | 10 |
| H/d of wear on compliant days ($\geq 10$h) | | | | | | |
|  | Mean (SD) | 12.1 (1.3) | 11.8 (1.4) | 12.1 (1.3) | 12.2 (1.3) | 12.2 (1.3) |
|  | Median | 12.0 | 11.5 | 12.0 | 12.1 | 12.2 |
| Passive trips/day |  |  |  |  |  |  |
|  | Mean (SD) | 1.8 (1.2) | 0.3 (0.3) | 1.4 (0.3) | 2.2 (0.2) | 3.5 (0.7) |
|  | Median | 1.8 | 0.3 | 1.4 | 2.2 | 3.3 |

### Strengths and limitations

To our knowledge, MacM3 is the **first longitudinal study designed to understand trajectories of mobility decline in a generalizable sample of community-dwelling older adults using a body-worn monitoring system that integrates GNSS and accelerometry**. The main strengths of the MacM3 study include its repeated measures and extended follow-up period, its innovative approach to mobility monitoring with advanced wearable technologies, the richness of the health and mobility data collected, and the cohort’s high participation and retention rate to date.

Although the use of wearable technology to measure real-world mobility holds great promise for improving our understanding of healthy aging, there are several challenges in this field. There is wide variability in the protocols used and parameters reported from wearable technology, with few studies evaluating their validity.^24,97,98^ Wear location of a sensor, for example, is critical to generating valid and sensitive outcomes.^99^ Additionally, much of the literature on wearable technology to date is from small cross-sectional studies, specific disease groups, or single sensor designs that can limit accuracy, generalizability and precision in the study of mobility in older adults.^100^ In recognition of some of these issues, a large European consortium—Mobilise-D— was formed to validate digital mobility measures for health and regulatory agencies. Mobilise-D is conducting a series of clinical validation studies in four specific disease cohorts (each *n* = 600) focusing on accelerometry-derived mobility measures such as gait speed.^24^ Our work complements that of Mobilise-D, as MacM3 will be the first cohort dedicated to evaluating digital mobility measures in large sample of community-dwelling older adults. Additionally, based on our extensive pilot testing,^27^ we have innovated a novel multi-sensor, multi-nodal approach that combines GNSS and accelerometry to optimize the potential for identifying early changes in mobility that could inform prevention and early intervention strategies from health, environmental, and social perspectives. This is a significant step forward in the study of real-world mobility in community-dwelling older adults, as GPS technology can provide an understanding of spatial properties of mobility patterns and environmental influences but lacks the motion and temporal fidelity to provide details of movement or walking. In contrast, accelerometry data provides details of movement and walking kinematics with continuous millisecond resolution but lacks spatial context. By temporally aligning accelerometry and GPS data, we leverage their complementary strengths to enhance the collection and interpretation of comprehensive mobility data, particularly for understanding environment-movement interactions. Furthermore, our cohort’s size, diversity (race, ethnicity, income, and ability), and range of health conditions and mobility levels will allow us to validate digital mobility measures with broad applicability for early risk assessment and intervention in community-dwelling older adults.

The main limitations of MacM3 include our convenience sample with recruitment from only 2 sites in Ontario, Canada, with a large proportion of individuals with post-secondary education and higher income levels. Our sample also consists of more females than males. The gender imbalance may similarly affect the generalizability of the findings, as males and females may have different patterns of mobility, health outcomes and health utilization. Another limitation is that wearing the thigh device was optional in this study, leading to the absence of key gait and posture-related behavior data for participants who chose not to wear it. Nevertheless, this limitation is mitigated by the high percentage of participants who agreed to wear the device, maintaining the overall usefulness of the dataset for studying gait and posture.

### Future Plans

Our future research will focus on leveraging the rich, longitudinal, multi-sensor data from MacM3 to develop predictive models of mobility decline. These models will enable early identification of individuals at risk of developing mobility limitations, guiding targeted interventions to prevent or mitigate disability. A key goal is to explore the integration of wearable-derived mobility metrics with clinical and environmental factors to personalize strategies for promoting functional independence.

Opportunities for extending the follow-up in MacM3 will also be sought to enhance the usefulness of the data platform and allow for rigorous validation of mobility measures obtained through multi-sensor wearable technology for monitoring and predicting long-term health outcomes in older adults.

Another critical avenue involves advancing the analytic capabilities of our open-source data processing pipeline, enhancing the fusion of GPS and IMU data to investigate the interaction between mobility behaviors and environmental contexts. By incorporating machine learning techniques and stakeholder input, we aim to translate these innovations into actionable tools for clinicians, caregivers, and older adults themselves. Ultimately, these efforts will contribute to developing scalable solutions that support aging-in-place, improve health outcomes, and enhance quality of life for aging populations.

## Collaboration

We encourage collaboration on projects that align with the goals of our study, particularly those focused on mobility and healthy aging. The dataset includes wearable sensor data (accelerometry and GPS), questionnaire responses, demographic and health information, as well as data from physical assessments. Researchers interested in accessing the data will be required to submit a proposal outlining their intended use, which will be reviewed by the study team. Data will be shared in a de-identified format whenever possible. However, certain restrictions may apply—particularly for GPS data—due to the sensitive nature of location information. Access to such data will require additional justification and safeguards to protect participant confidentiality.

## MacM3 Investigators (Collaborators)

Jamal Deen, Electrical and Computer Engineering Department, McMaster University; Vanina Dal Bello-Haas, McMaster University; Rebecca Ganann, School of Nursing, McMaster University; Jack Guralnik, Department of Epidemiology and Public Health, University of Maryland School of Medicine; Lori Letts, School of Rehabilitation Science, McMaster University; Sarrah Lal, Faculty of Health Sciences, McMaster University; Stuart Phillips, Department of Kinesiology, McMaster University, McMaster University; Nigar Sekercioglu, Health Research Methods, Evidence, and Impact, McMaster University; Kathryn Sibley, Max Rady College of Medicine, University of Manitoba.

## Authors Contribution

**MKB, JR, AK, KBN, DMS, PR, QF, PG, PDM, and MZ conceived the study**. MKB, JR, AK, PDM, KBN, DMS, PR, QF, PM, MZ, JD, VDBH, RG, JG, LL, SL, SP, NS and KS contributed to the study design. MKB, RK, CC and SO were responsible for the study planning and data collection. MKB, RK, CC, WM, KVO and KB were responsible for developing the protocol for analyzing the wearables data. MKB, RK, JM, and TR were responsible for the analysis. MKB, RK and TR were responsible for writing the initial draft of the manuscript, and subsequent drafts were reviewed by all authors listed. All authors had input on the interpretation and reporting of study findings. All authors provided approval for the published version of this manuscript. MKB is the guarantor of the work and accepts full responsibility for the integrity of the data and the accuracy of the analysis.

## Acknowledgements

The authors would like to thank the study coordinators, Tara Noble and Lisette Machado, and the research assistants—Kathleen Romanoski, Jasmyn Stoffers, Rhea Sinha, Mae Vidal, Shalini Jaisankar Sumathi and Catherine Deng — for their support throughout data collection and participant coordination.

## Competing interests

All authors have no competing interest to declare.

### Ethics approval

This study was approved by the Hamilton Integrated Research Ethics Board (HiREB), McMaster University (Project number: 13905).

## Data availability statement

Access to MacM3 data is currently unavailable to other researchers. We are working on developing processes to make de-identified data available to qualified researchers for approved analyses, following the review and approval of a study proposal, data use agreement, and ethical approval. For more information, please contact the corresponding author at.

