## Supplemental Table 2 for "Cohort Profile: Baseline characteristics and design of the McMaster Monitoring My Mobility (MacM3) Study, a prospective digital mobility cohort of community-dwelling older Canadians from Southern Ontario"

Supplement 1. Selected baseline characteristics of the MacM3 participants.

|  | | Total | Hamilton site | | Dixon Hall site | |
| --- | --- | --- | --- | --- | --- | --- |
|  |  |  | Males  *n* (%) | Females  *n* (%) | Males  *n* (%) | Females  *n* (%) |
| Sample size, *n* (%) | | 1555 | 408 (33.7) | 802 (66.3) | 83 (24.0) | 262 (76.0) |
| Cultural backgrounds, *n* (%) *(participants could select multiple cultural identities)* | English  Scottish  Irish  Canadian  German  Chinese  French  Italian  Polish  Dutch  Other (Ukrainian, Jewish, Welsh, South Asian, Norwegian, Swedish, Portuguese, Metis, North American Indian, other) | 622 (40.0)  391 (25.1)  375 (24.1)  226 (14.5)  157 (10.1)  109 (7.0)  104 (6.7)  83 (5.3)  83 (5.3)  67 (4.3)  478 (30.7) | 165 (40.4)  99 (24.3)  109 (26.7)  70 (17.2)  48 (11.8)  1 (0.2)  29 (7.1)  23 (5.6)  22 (5.4)  19 (4.7)  131 (32.1) | 372 (46.4)  237 (29.6)  211 (26.3)  143 (17.8)  80 (10.0)  5 (0.6)  55 (6.9)  52 (6.5)  38 (4.7)  38 (4.7)  226 (28.2) | 17 (20.5)  14 (16.9)  5 (6.0)  4 (4.8)  6 (7.2)  32 (38.6)  1 (1.2)  1 (1.2)  8 (9.6)  2 (2.4)  21 (25.3) | 68 (26.0)  41 (15.6)  50 (19.1)  9 (3.4)  23 (8.8)  71 (27.1)  19 (7.3)  7 (2.7)  15 (5.7)  8 (3.1)  100 (38.2) |
| Comorbidities, *n* (%) | None | 54 (3.5) | 21 (5.1) | 21 (2.6) | 2 (2.4) | 10 (3.8) |
|  | < 3 | 522 (33.6) | 126 (30.9) | 270 (33.7) | 30 (36.1) | 96 (36.7) |
| Common conditions, *n* (%) | ≥ 3  Cataracts  Osteoarthritis  Hypertension  Back Pain  Osteoporosis  Cancer  Diabetes  Depression | 979 (62.9)  684 (44.0)  652 (41.9)  568 (36.5)  268 (17.2)  247 (15.9)  246 (15.8)  174 (11.2)  170 (10.9) | 261 (64.0)  158 (38.7)  145 (35.5)  188 (46.1)  68 (16.7)  11 (2.7)  79 (19.4)  73 (17.9)  30 (7.4) | 511 (63.7)  359 (44.8)  381 (47.5)  262 (32.7)  141 (17.6)  151 (18.8)  98 (12.2)  68 (8.5)  94 (11.7) | 51 (61.5)  40 (48.2)  24 (28.9)  36 (43.4)  15 (18.1)  11 (13.3)  17 (20.5)  15 (18.1)  5 (6.0) | 156 (59.5)  127 (48.5)  102 (38.9)  82 (31.3)  44 (16.8)  74 (28.2)  52 (19.8)  18 (6.9)  41 (15.6) |
| Prescribed medication, *n* (%) | Yes | 1,261 (81.2) | 355 (87.5) | 652 (81.4) | 71 (85.5) | 183 (69.8) |
| Number of prescribed medications, *n* (%) | No  0  1  2  3  4  5 or more | 291 (18.8)  291 (18.7)  312 (20.1)  265 (17.0)  243 (15.6)  169 (10.9)  272 (17.5) | 51 (12.5)  51 (12.5)  56 (13.7)  62 (15.2)  66 (16.2)  53 (13.0)  118 (28.9) | 149 (18.6)  149 (18.6)  169 (21.1)  143 (17.8)  132 (16.5)  88 (11.0)  120 (15.0) | 12 (14.5)  12 (14.5)  13 (15.7)  15 (18.1)  18 (21.7)  8 (9.6)  17 (20.5) | 79 (30.2)  79 (30.2)  74 (28.2)  45 (17.2)  27 (10.3)  20 (7.6)  17 (6.5) |
| Self-rated general health, *n* (%) | Excellent | 258 (16.6) | 58 (14.2) | 134 (16.7) | 7 (8.4) | 59 (22.5) |
|  | Very good | 687 (44.2) | 184 (45.1) | 368 (45.9) | 27 (32.5) | 108 (41.2) |
|  | Good | 474 (30.5) | 129 (31.6) | 249 (31.0) | 36 (43.4) | 60 (22.9) |
|  | Fair | 123 (7.9) | 34 (8.3) | 49 (6.1) | 11 (13.3) | 29 (11.1) |
|  | Poor  Don’t know/No answer | 10 (0.6)  2 (0.1) | 3 (0.7)  - | 1 (0.1)  1 (0.1) | 2 (2.4)  - | 4 (1.5)  1 (0.4) |
| Self-rated mental health, *n* (%) | Excellent | 286 (18.4) | 77 (18.9) | 129 (16.1) | 18 (21.7) | 62 (23.8) |
|  | Very good | 647 (41.6) | 177 (43.4) | 332 (41.4) | 26 (31.3) | 112 (42.7) |
|  | Good | 515 (33.1) | 130 (31.9) | 296 (36.9) | 28 (33.7) | 61 (23.3) |
|  | Fair | 89 (5.7) | 21 (5.1) | 41 (5.1) | 7 (8.4) | 20 (7.6) |
|  | Poor  Don’t know/No answer | 15 (1.0)  2 (0.1) | 3 (0.7)  - | 4 (0.5)  - | 4 (4.8)  - | 4 (1.5)  2 (0.8) |
